# Centers for Mendelian Genomics: A decade of facilitating gene discovery

**DOI:** 10.1101/2021.08.24.21261656

**Authors:** Samantha M. Baxter, Jennifer E. Posey, Nicole J. Lake, Nara Sobreira, Jessica X. Chong, Steven Buyske, Elizabeth E. Blue, Lisa H. Chadwick, Zeynep H. Coban-Akdemir, Kimberly F. Doheny, Colleen P. Davis, Monkol Lek, Christopher Wellington, Shalini N. Jhangiani, Mark Gerstein, Richard A. Gibbs, Richard P. Lifton, Daniel G. MacArthur, Tara C. Matise, James R. Lupski, David Valle, Michael J. Bamshad, Ada Hamosh, Shrikant Mane, Deborah A. Nickerson, Centers for Mendelian Genomics Consortium, Heidi L. Rehm, Anne O’Donnell-Luria

**Author notes:** Corresponding authors Contact information for corresponding authors: Ann O’Donnell-Luria, Heidi Rehm, Samantha Baxter.

## Abstract

Mendelian disease genomic research has undergone a massive transformation over the last decade. With increasing availability of exome and genome sequencing, the role of Mendelian research has expanded beyond data collection, sequencing, and analysis to worldwide data sharing and collaboration. Over the last 10 years, the NIH-supported Centers for Mendelian Genomics (CMGs) have played a major role in this research and clinical evolution. We highlight the cumulative gene discoveries facilitated by the program, biomedical research leveraged by the approach, and the larger impact on the research community. Mendelian genomic research extends beyond generating lists of gene-phenotype relationships, it includes developing tools, training the larger community to use these tools and approaches, and facilitating collaboration through data sharing. Thus, the CMGs have also focused on creating resources, tools, and training for the larger community to foster the understanding of genes and genome variation. The CMGs have participated in a wide range of data sharing activities, including deposition of all eligible CMG data into AnVIL (NHGRI’s Genomic Data Science Analysis, Visualization, and Informatics Lab-Space), sharing candidate genes through Matchmaker Exchange (MME) and the CMG website, and sharing variants in Geno2MP and VariantMatcher. The research genomics output remains exploratory with evidence that thousands of disease genes, in which variant alleles contribute to disease, remain undiscovered, and many patients with rare disease remain molecularly undiagnosed. Strengthening communication between research and clinical labs, continued development and sharing of knowledge and tools required for solving previously unsolved cases, and improving access to data sets, including high-quality metadata, are all required to continue to advance Mendelian genomics research and continue to leverage the Human Genome Project for basic biomedical science research and clinical utility.

## Introduction

The completion of the first human reference genome in 2001, complemented by the rapid development of next-generation sequencing (NGS), brought about a paradigm shift in the field of human genetics and Mendelian disease research. Candidate-gene sequencing, positional cloning, and physical mapping of genes were rapidly replaced when NGS technologies enabled routine examination of all human coding regions in a single analysis. By 2010, it was clear that NGS methods, particularly exome sequencing (ES), offered a robust approach to identify candidate novel disease genes and molecular diagnoses.^1–5^ With these advancements, it was now possible to study Mendelian conditions in a rapid and cost-effective manner. High-throughput sequencing approaches lent themselves to coordination through national programs to allow the efficient generation of high-quality genomic data analyzed through rigorous sustainable pipelines. Effectively finding causes for the rarest of diseases requires increasing the reach of genomic research through training more clinicians and researchers in genomic analysis and a robust infrastructure that supports data sharing and interdisciplinary collaborative research.

With a focus on identifying the causative variants in the genes responsible for all Mendelian phenotypes, several national and international efforts were quickly developed, and included collaborative efforts such as the FORGE Canada Consortium,^6^ Deciphering Developmental Disorders in the UK study,^7^ Undiagnosed Diseases Network in the US^8^, and the International Rare Diseases Research Consortium (IRDiRC).^9^ In 2011, the National Human Genome Research Institute (NHGRI) within the US’s National Institutes of Health (NIH) established the Centers for Mendelian Genomics (CMGs), with additional support from the National Heart Lung and Blood Institute (NHLBI) and later the National Eye Institute (NEI). The need for common infrastructure, workflows and methods development across all disease areas provided the rationale for a centralized CMG structure that could support national and international collaborative efforts with clinicians and researchers who might not otherwise have the necessary resources or environments to engage in genomic research -- thus, truly opening the door to the study of all Mendelian conditions. The goal of the CMG program was to accelerate the identification of genetic variation underlying Mendelian conditions leveraging genome-wide NGS technologies, and to disseminate discoveries, approaches, and technology broadly to drive discovery worldwide (described in Bamshad *et al*. 2012).^10^ The consortium initially consisted of three centers: the Baylor-Hopkins CMG, the University of Washington CMG, and the Yale CMG. A fourth center, the Broad Institute of MIT and Harvard CMG, was added during the second phase of the consortium. Now in the final year of funding, we reflect on the data generated, accomplishments, lessons learned, and remaining challenges from a decade of gene discovery by the CMGs. We highlight the final achievements of the CMGs through discoveries, data sharing, tools, and impact on the community and discuss needed next steps to further synergize genetics and genomics. We have learned much from the CMGs, and while the CMGs are coming to an end, a new NHGRI-funded program, the Mendelian Genomics Research Consortium (MGRC), will begin in 2021. The MGRC’s mandate is to sequence, identify, and validate disease-contributing genes and variants in families for whom current approaches have failed to find a molecular diagnosis, essentially developing approaches to ‘solve the unsolved.’

### The importance of collaboration in Mendelian gene discovery

The rarity and diversity of Mendelian diseases require that we cast a broad net across the human population to meet our goal of complete enumeration of Mendelian disease genes. Accordingly, the four CMGs collaborated directly with 2,283 researchers during the last decade. Through extended collaborations and publications, the CMGs worked with 11,771 researchers across 2,773 institutions in 90 countries. Building this vast collaborative network required global outreach efforts including advertisements in the *American Journal of Medical Genetics*, seminars at conferences, educational courses, ‘boots on the ground’-style recruitment, and word of mouth from early collaborators.

Collaborators brought DNA samples from deeply phenotyped, unsolved families affected by rare disease consented for genomic studies and data sharing; CMGs provided sequencing, data processing, and data sharing, and analysis was done in collaboration through joint distributive research efforts. While collaboration styles varied based on the preferences of the individual CMGs and collaborators, often the CMGs would perform initial genomic analyses to prioritize a short list of high priority candidate variants for each proband/family that could be discussed in consultation with the collaborator. Collaborators typically pursued the variants of interest to gather additional cases, performed functional studies, synthesized the science, and wrote up the findings, often with resources and support from the CMGs.

Most successful discoveries required collaboration that extended beyond one CMG and a single collaborator. In an analysis of the CMG’s gene-discovery publications, we found that more than 90% involved more than one institution contributing cases to the publication, highlighting the fundamental role of collaboration in solving Mendelian disease. These connections are often made via data sharing platforms and activities, which have become a critical component to gene discovery.^11^

In 10 years, the CMGs have contributed to 961 manuscripts by providing sequencing data, analysis, methods development, along with training in genomic analysis. The CMGs have generated 75,573 exomes, 3,876 genomes, 714 transcriptomes through RNA-seq, and 385 methylation arrays across 28,991 families (Figure 1). Samples that have been pre-screened for known causes of disease are prioritized by the CMGs, though as sequencing prices fell over the course of the program and the approach of family-based genomics became better defined, this became less pragmatic, particularly for samples and investigators from under-resourced countries. Prior data freeze assessments of the first and second phases of the Consortium highlight both the scale and breadth of Mendelian phenotypes studied, as well as the international and collaborative impact of the CMGs. ^12, 13^

**Figure 1.**
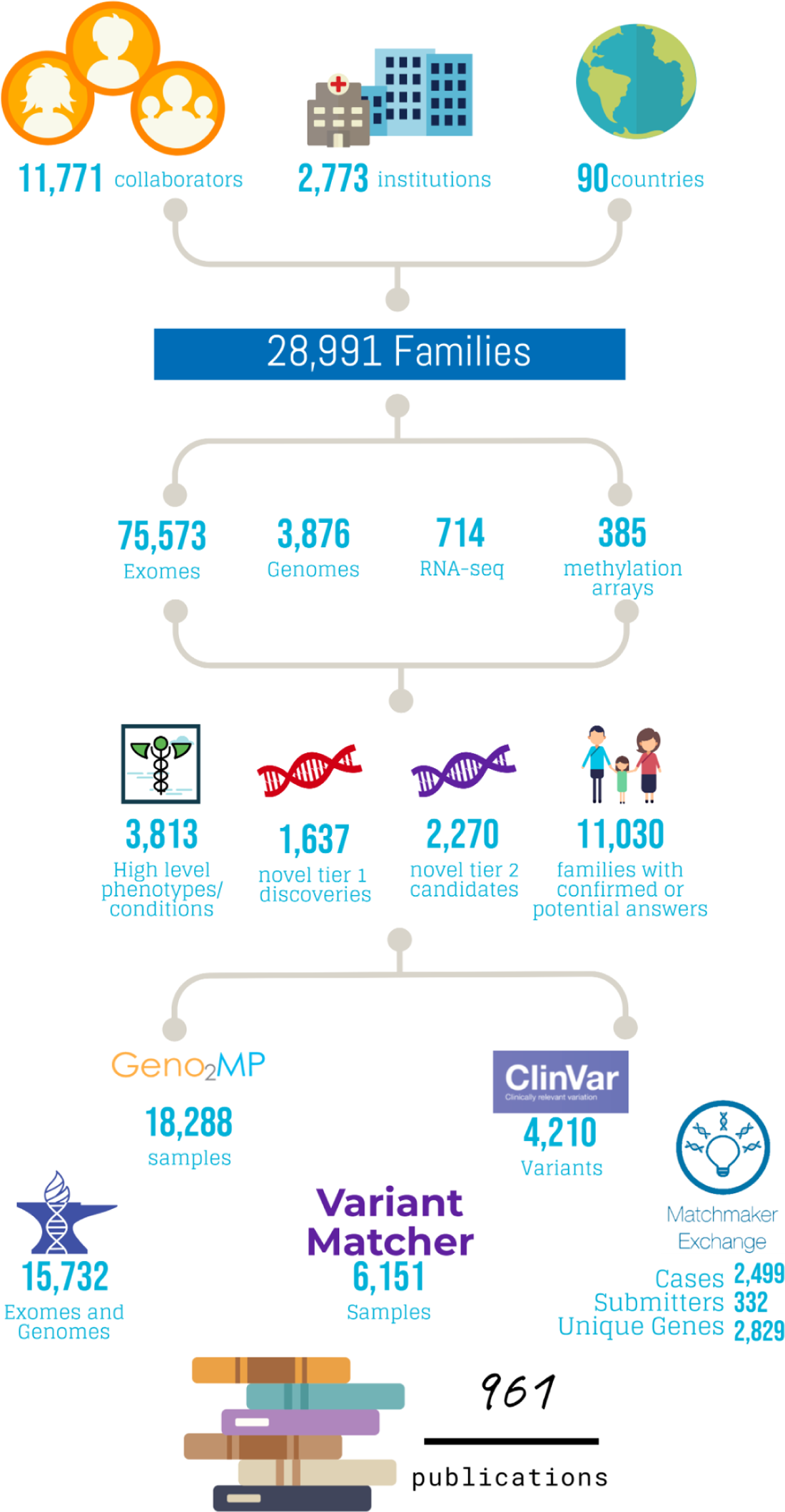
Overview of CMG by numbers. A high level summary of activities performed by the CMGs including the number of collaborators, number of kindreds, volume of testing performed, discovery rates, and data sharing metrics.

### Track record in discovering gene-disease relationships

Remarkably, the rate of CMG discovery has continued on a fairly constant trajectory over the decade of the program.^12, 13^ We distinguish the known phenotypes from the novel phenotypes using OMIM’s Phenotype MIM IDs. The CMGs have identified 1,637 new disease-gene relationships (termed tier 1 for multiple affected kindreds identified for a gene-disease relationship or very strong functional data supporting the relationship) (172/year) and 2,270 candidates (termed tier 2) (239/year) candidates over the course of the program (Table 1). These Tier 2 candidates are strong enough to enter the gene in Matchmaker Exchange, often having some data in the literature implicating the gene for the phenotype, but further data is needed such as additional unrelated probands with an overlapping phenotype, model organism, or other functional studies.

**Table 1.**
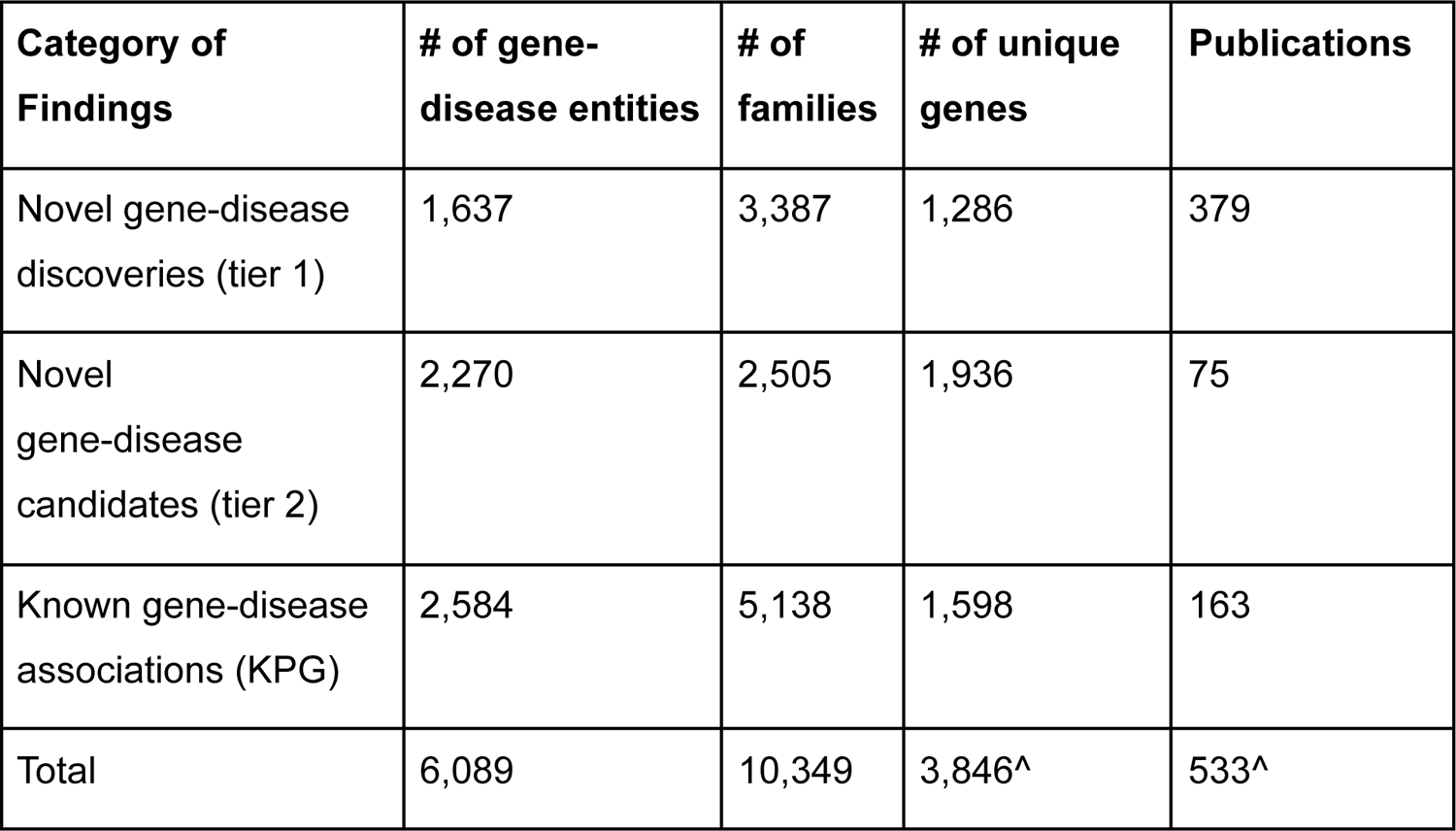
Breakdown of CMG findings by category ^total for unique genes and publications is not equal to the sum of the rows above because genes can have multiple disease associations, with varying certainty of associations, and publications can be duplicated across categories

| **Category of Findings** | **# of gene-disease entities** | **# of families** | **# of unique genes** | **Publications** |
| --- | --- | --- | --- | --- |
| Novel gene-disease discoveries (tier 1) | 1,637 | 3,387 | 1,286 | 379 |
| Novel gene-disease candidates (tier 2) | 2,270 | 2,505 | 1,936 | 75 |
| Known gene-disease associations (KPG) | 2,584 | 5,138 | 1,598 | 163 |
| Total | 6,089 | 10,349 | 3,846<sup>^</sup> | 533<sup>^</sup> |

This continued rate of discovery tells us that there are still more Mendelian genes to be found, and that ongoing sequencing efforts should continue to be fruitful. Details of each novel gene-disease association are collected by the CMGs, including information about the phenotypes being investigated, the variant details, and whether the discoveries being reported are for genes with no existing disease relationships or if the new discovery represents a phenotype expansion for the gene (supplemental table 1).

There are intriguing observations and interesting trends in the success of gene-disease discovery efforts across organ systems, although we note the caveat that efforts have not been applied evenly. Much effort has focused on neurodevelopmental conditions and syndromic disorders where there has been a continued high rate of gene-disease discovery (Figure 2). For cases where only a single system was impacted, disorders of blood and blood-forming tissues, metabolism/homeostasis, the immune system, integument, and the nervous system have the highest rates of novel findings. When multiple systems are involved, involvement of the skeletal system or connective tissue has the highest rate of findings (including known gene-disease associations, novel gene-disease discoveries and candidates). Skeletal abnormalities or involvement of the immune system had the highest rate of novel discovery. For ear, eye, kidney, connective tissue and muscle phenotypes, some new genes have been identified but many cases were solved with variants in genes that have known gene-phenotype relationships in OMIM. Solving many of these cases required going beyond standard analysis approaches or additional omics approaches, highlighting the impact of CMG sequencing and variant detection.^14–16^

**Figure 2.**
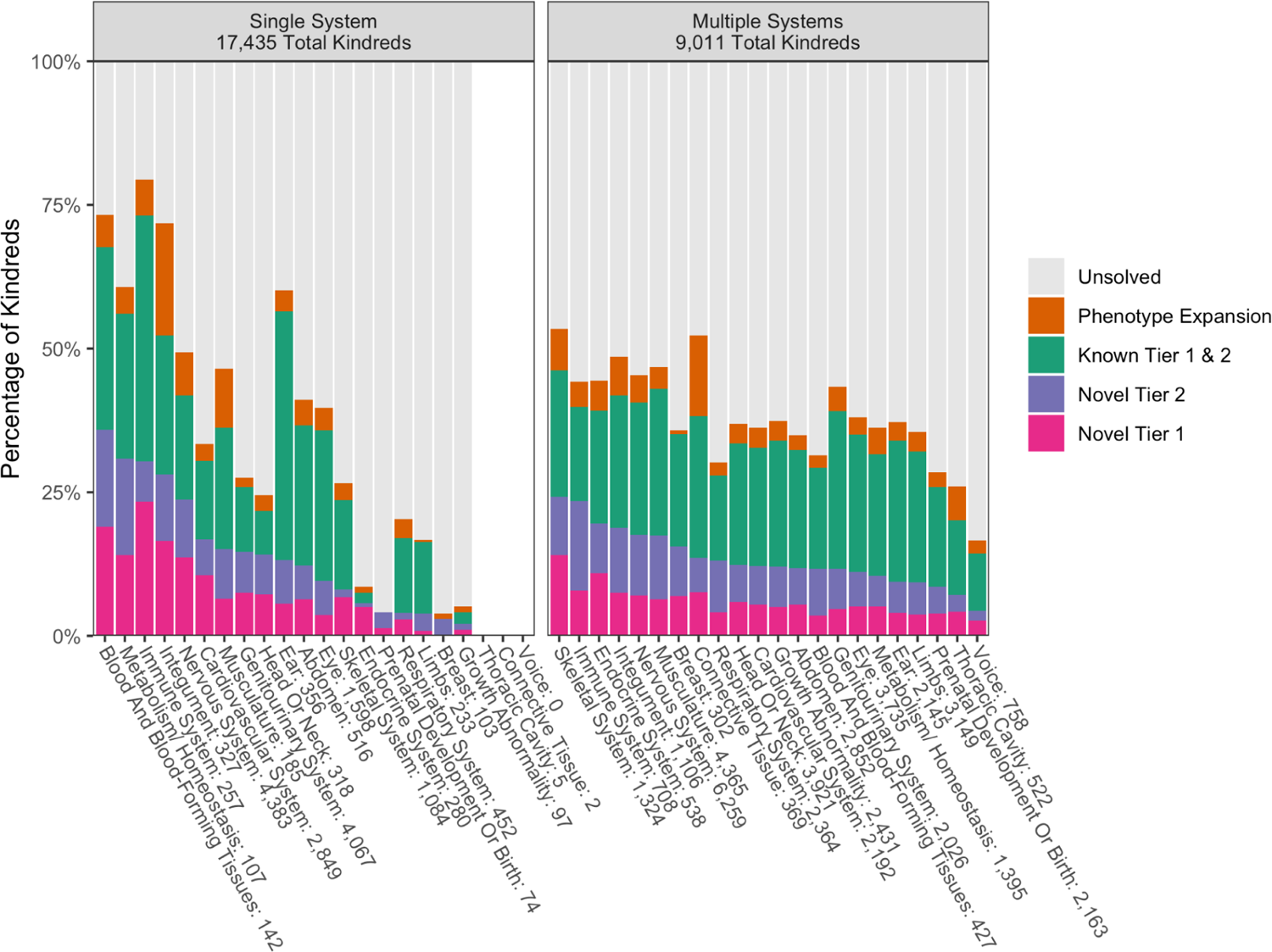
Solve and discovery rates by high level HPO category. Kindreds were categorized as having phenotypes in one HPO high-level system or multiple high-level systems. From there the solve and discovery types were analyzed for each system. There were 494 kindreds in our CMG cohort with no HPO terms available, and therefore they were not able to be included in this analysis. Systems with fewer than 10 kindreds were noted but excluded.

The CMGs have contributed to the discovery of 778 phenotypic expansions associated with previously established disease genes. Such discoveries represent an important contribution to both the research and clinical fields, as the full phenotypic spectrum of a Mendelian disease, or the set of phenotypes associated with a genomic locus, may not be fully revealed at the time of the initial disease gene discovery. Indeed, approximately 961 genes, or 24% of genes implicated in Mendelian disease, now have more than one phenotype associated with the gene.^17, 18^ The rate of disease gene discovery continues to outpace that of phenotype expansion, highlighting the continued value of Mendelian disease gene discovery. Many of the CMG discoveries are also uncovering molecular and biological processes that can inform therapeutic intervention or management, ranging from existing medications known to impact a particular metabolic pathway or ion channel, to avoidance of certain medications, to bona fide molecular interventions.^19–23^

### Impact on the rare disease community

To advance discovery of the underlying basis of Mendelian disease, interactions with clinical laboratories are important, both to relay discoveries more rapidly and efficiently than the peer review publication process allows, as well as to access primary evidence from patient testing. However, navigating the clinical research boundaries can be challenging. Many genes discovered and published through traditional research still lack the evidence required to definitively establish a role in disease and enable the molecular diagnosis of patient symptomatology.^24, 25^ Including genes with limited evidence on clinical testing panels increases the number of variants of uncertain significance (VUS) on patient’s test results without increasing yield, which has led many labs to set a threshold of evidence for genes to be included on clinical test reports. Giving patients uncertain results can distract them from pursuing other causes of disease; yet in other circumstances, empower them to engage in the research process. Thus, researchers, clinical labs, and patients must work together to share primary evidence, build the clinical genomic knowledgebase, and ensure rapid translation of discoveries to patient care.

Thus far, the CMGs have contributed to 419 publications, describing novel gene-disease discoveries and candidate gene-disease associations, while another 163 articles have added to the understanding of known gene-disease associations. To date, 23.2% (379) of the 1,637 Tier 1 novel gene-disease discoveries have been published in peer-reviewed journals. Many of the CMG discoveries are still in the process of gathering cases, functional data, or going through the peer review publication process.

To date, 836 (65.0%) of the CMGs unique novel gene-disease discoveries have a Pathogenic/Likely Pathogenic variant, submitted by a clinical laboratory, in ClinVar. Of these 836 genes, 496 genes (59.3%) have not yet been published by the CMGs in peer-review journals, showing that these genes have successfully moved from research to the clinic and reflecting the success of data sharing practices beyond the peer-review publication process alone.

While the primary goal of the CMGs was gene discovery, their impact on the genetic community goes beyond establishing or clarifying gene-disease relationships. Through community outreach, training, and data sharing, CMG provided students, clinician-scientists, and investigators with the training and tools to discover new Mendelian disease genes. The CMGs provided educational and networking opportunities by hosting in-person courses, attended by over 300 analysts and researchers. Since 2013, the University of Washington CMG has hosted a week-long Mendelian Data Analysis Workshop on strategies and tools for solving Mendelian conditions using next generation sequence data. The Broad CMG has offered “Interpreting Genomes for Rare Disease” since 2017, which includes both lectures and breakout sessions for hands-on training on genomic filtration and gene/variant curation. Lectures were recorded and posted so the community has ongoing access (cmg.broadinstitute.org/course-offering). The Baylor-Hopkins CMG has incorporated exome analysis training into the McKusick-Short Course in Human and Mammalian Genetics training over 1500 students in in-person (through 2019) and virtual (2020 and 2021) workshops. These have also been held in Brazil and Chile. A special week-long Genomics course is incorporated into the medical school curriculum at Johns Hopkins and will be developed into an on-line training module. The CMGs have enabled researchers and clinicians investigating rare Mendelian diseases in the US and around the world to access gene discovery techniques, including those in countries where access to research opportunities is limited, such as the Democratic Republic of the Congo,^26^ South Africa, Kenya, Egypt,^27, 28^ Iraq,^29^ Chile ^30, 31^, Turkey,^32–34^ and Lithuania. For some, this has involved training opportunities within a CMG-affiliated laboratory, while for others learning happened through collaborative meetings to discuss analysis results on teleconferences.

To better understand the impact of the CMG program, we surveyed CMG collaborators in early 2021. A total of 206 responses were collected, including collaborators from basic research (37%), clinical research (33%), clinical practice (24%), and diagnostic laboratory (6%) areas. Most were senior investigators with >10 years of experience (63%), while junior faculty with <5 years (9%) and 5-10 years of experience (20%) were also well-represented; some were not in faculty positions (8%). Seventy six percent reported that they were more likely to share data as a result of their work with the CMGs, while the remainder were as likely as before. Seventy percent also reported starting new collaborations due to their work with a CMG. Most collaborators (69%) clinically confirm and return results identified through the CMG to the rare disease subjects and families, but several groups were unable to because of insufficient resources.

### Sharing data to improve rare disease diagnosis

Given the genetic heterogeneity and rarity of Mendelian disease and the diversity of our species, rapid and international data sharing is critical to build sufficient evidence to unambiguously identify novel gene-disease relationships. Many of the discoveries made by the CMGs were only possible because of collaborations, which were necessary for obtaining enough samples to convincingly associate a gene with a given phenotype. Data sharing is another key element in helping researchers with related cases find each other and work together.

Approaches that share only the results or top candidates from sequencing studies are inadequate to maximize discovery rates, as not all pathogenic variants are equally recognizable. For example, when a *de novo* predicted loss-of-function (pLOF) variant is identified, that researcher will want to look carefully at the phenotypes of any individuals with other pLOF or *in silico*-predicted damaging missense variants even if the gene was not flagged. Mendelian disease investigators therefore should design the infrastructure of each study in such a way that enables easy sharing of both the genomic data and accompanying metadata.

The CMGs have committed to rapid and extensive data sharing (table 2), developing new platforms accessible to the global community, and requiring participation by all CMG collaborators to the benefit of other researchers and rare disease patients. The CMG shares data using a variety of tools that support one of three primary modes of sharing: connecting two parties that each have a predefined candidate gene (*two-sided matching)* regardless of whether the variants in those genes are the same or different; allowing one party to query another party’s primary data for variants of interest or the presence of a gene with a type of variant (pLOF, biallelic rare variants, etc) (*one-sided matching*); or allowing credentialled researchers who apply for access to the data to directly analyze cohorts of read-level sequence data (shared via CRAM files) and detailed meta-data in structured files (Figure 1) to make discoveries in the absence of any identified gene candidates (*zero-sided matching*). By participating in multiple types of data sharing, data sequenced through the CMGs is made accessible to the broadest possible range of researchers and use-cases across the rare disease community.

**Table 2:**
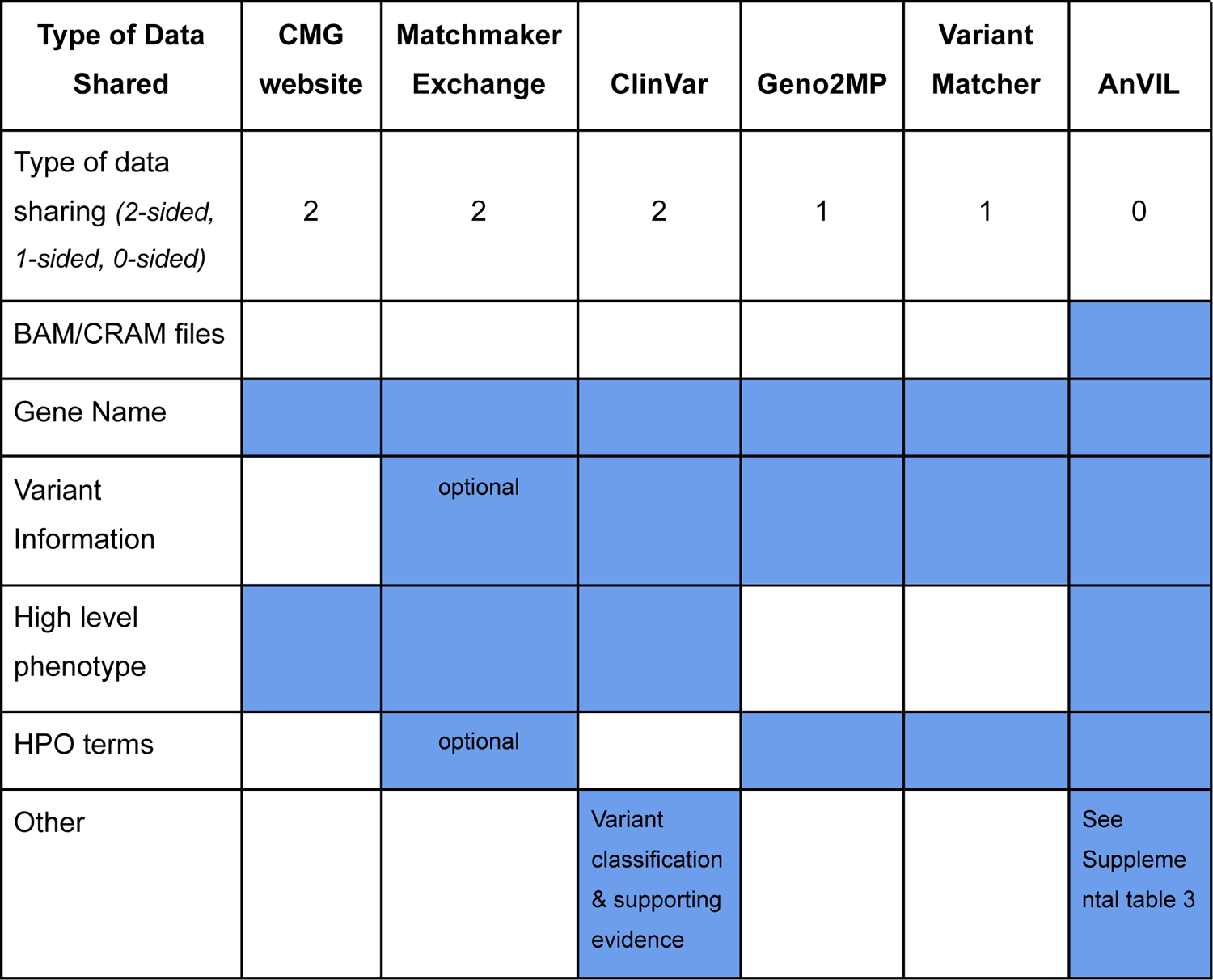
CMG Data Sharing. A breakdown for the various data sharing activities in which the CMGs participated. Each column is a different data sharing platform, and the rows are the types of data that the CMGs could share. The **blue boxes** indicate that the CMGs submitted that file or data type to the platform.

### Sharing for two-sided matching (both parties have gene candidates for their cases)

Two-sided matching historically occurred through parties listing and/or searching for genes of interest on websites, using search engines like google as the “matcher”, or “cold emailing” inquiries to colleagues. This method can be error prone, time-consuming, and lead to either overly inclusive results or limited results. However, websites like ClinVar or mendelian.org allow for data to be shared quickly and openly to anyone with internet access. More recently, informatics has been used to create genomics matchmaking tools which allow matching based on gene and/or phenotype. The CMGs have contributed to both website-based and informatics-enabled matching.

#### CMG Website

The coordinating center of the NHGRI Genome Sequencing Program has created a website to disseminate information arising from the CMG program (http://mendelian.org/). In an effort to release as many of our candidate gene-phenotype relationships as possible, this site hosts a searchable list of phenotypes under investigation, as well as candidate gene-phenotype relationships that meet Tier 1 or Tier 2 criteria (http://mendelian.org/phenotypes-genes). This site also maintains a list of all publications that acknowledge support from one of the CMG centers (http://mendelian.org/publications).

#### Matchmaker Exchange

Matchmaker Exchange (MME) plays a critical role in facilitating the aggregation of individual cases with variants in a given candidate gene. MME is a federated network designed to connect databases of gene candidates and phenotypic data via a common API. It allows for computational ranking of matches by phenotype, and enables collaborators to connect for follow-up and more detailed manual comparisons of phenotypes.^35^ In addition to committing to sharing candidate genes to MME, CMGs have also contributed to this platform by building 3 of the 8 current nodes (GeneMatcher, MyGene2, and *seqr;* table 3).

**Table 3:** CMG gene candidate sharing through Matchmaker Exchange (MME). This is a summary of the data submitted to the MME through CMG-developed nodes. The number of cases, submitters, and unique genes that were contributed to the MME are broken down into CMG and non-CMG cases, as non-CMG participants are still able to use the CMG-developed nodes to share on MME.

| MME Node | CMG Developer | CMG submitter | Case in MME |  | Submitters |  | Unique Genes |  |
| --- | --- | --- | --- | --- | --- | --- | --- | --- |
|  |  |  | CMG | non - CMG | CMG | non - CMG | CMG | non - CMG |
| GeneMatcher | BH | BH | 343 | 55,210 | 99 | 11,790 | 817 | 12,964 |
|  |  | Yale | 270 | 0 | 36 | 0 | 276 | 0 |
| MyGene2 | UW | UW | 871 | 599 | 119 | 125 | 635 | 563 |
| *seqr* | Broad | Broad | 1,015 | 70 | 78 | 13 | 1,101 | 81 |

**GeneMatcher** (https://genematcher.org), developed by the Baylor-Hopkins CMG, only requires submission of a gene name, but also permits the inclusion of phenotypic features, conditions, and variants to improve the utility and specificity of matching.^36^ Any researcher, clinician, patient or family may register with GeneMatcher and submit candidates. GeneMatcher notifies submitters immediately if matches are identified within GeneMatcher, and can also return matches from connected MME nodes. Model organism researchers can also submit genes to GeneMatcher; there are currently 270 submissions that fall into this category, a majority of which are from mouse models.

**MyGene2** (https://mygene2.org), developed by the University of Washington CMG, is a website that enables families, clinicians, and researchers to openly and publicly share genetic information such as candidate genes and variants alongside phenotype with the goal of facilitating research into rare genetic conditions. ^37^ Candidate gene/variant and phenotype data can be browsed and searched by the public and are available for automated matching within MyGene2. Additionally, potential novel gene-disease relationships are shared through MME for matching beyond the MyGene2 node. MyGene2 is designed to be family-friendly so that all stakeholders, including families with rare conditions can join clinicians and researchers in learning from, contributing to, and even engaging in research through the site.

The ***seqr*** Matchmaker Exchange node (formerly known as *matchbox* ^38^; https://seqr.broadinstitute.org), developed by the Broad CMG, is directly integrated with the open source *seqr* genomic analysis web application. Users of the *seqr* application are able to submit gene name, variant details, and HPO terms to MME for discoveries made within *seqr*, to match with other MME nodes. The interface allows tracking of match outcomes and auto-generation of email text with case and variant details to contact potential matches.

#### ClinVar

In addition to facilitating novel gene-disease associations and increasing solve rates, data generated by the CMGs can be valuable for many other clinical and research uses, including clinical molecular diagnosis, characterizing the natural history of disease, and providing the foundational data to catalyze interventional and disease mitigation therapeutic strategies. To support these efforts, the CMGs have committed to sharing all published variants and supporting evidence to ClinVar, an open-source database that collects and reports variant and phenotype relationships. Since the inception of the program, the CMGs have together submitted over 4,200 variants, along with supporting evidence summaries, to ClinVar. These classifications can be used by diagnostic labs and other researchers to improve interpretation of variants and molecular diagnosis rates. The center-specific names and pages can be found in supplemental table 2.

### Sharing for one-sided matching (only one party has a gene candidate for their case and wishes to query other primary datasets to find a match that was not previously recognized by the other party)

While two-sided matching is helpful when both parties share a gene or phenotype of interest, many researchers have cases that don’t yet have an identified candidate gene. In addition to these unsolved cases without candidates, there remain many candidate genes without matches, highlighting the need to search databases of rare disease patients for additional copies of specific candidate variants or other similar potentially deleterious variants within the candidate genes. The CMGs have created two databases to support one-sided variant matching. These databases also help to exclude candidates, as the variant of interest may be found in unaffected relatives or well-phenotyped individuals with unrelated phenotypes with documented absence of the phenotype in question. Nevertheless, caution must be exercised in ‘exclusion’ based on nonpenetrance in a family member.

**Geno2MP (**Genotypes to Mendelian Phenotypes) was created by the University of Washington CMG to facilitate new gene discovery efforts and prioritization of candidate variants. It contains aggregate genotype data from >19,000 samples (>15 million rare variants with <2.5% frequency in gnomAD) sequenced by the UW, Broad, and Yale CMGs, which can be queried via the Geno2MP Browser (https://geno2mp.gs.washington.edu/). All variants are available for browsing on a gene page that includes information about the variant type and its phred-scaled CADD score.^39^ Each rare variant is linked to de-identified phenotypic information about the affected individuals and unaffected relatives who carry the variant. Geno2MP users may contact original submitters of the samples through the website and provide details about their own cases in order to pursue potential gene discovery matches.

**VariantMatcher**, created by the Baylor-Hopkins CMG, allows users to query rare (<1% allele frequency), non-synonymous variants from over 6,151 samples sequenced by the BHCMG for specific variants of interest. Users of the site must register and be approved by site administrators. If phenotypic features are submitted in combination with the variant in the query, the phenotype from the matched entry will be shared in a simultaneous email to the submitter and the matching investigator. If a match is not made, the queried coordinates with submitted phenotypes can be stored for future matching.

### Sharing for zero-sided matching (querying larger, combined datasets to identify novel gene-disease candidates in the absence of any prior identified gene candidate)

Novel methods and aggregation of larger datasets will be needed to solve cases that remain unsolved after thorough analysis by the CMGs. In the initial years of the program, cram files from the CMGs were shared through dbGaP (NIH Database of Genotypes and Phenotypes), but access for analysis required download and storage. The center-specific study accession numbers can be found in supplemental table 3. As genomic data grows in scale, the community needed solutions like NHGRI’s AnVIL, where the analysis tools can be brought directly to the data, and where researchers of varying computational skill levels are enabled to interrogate it. CMG data sharing has transitioned to AnVIL, where >60 terabytes of data are available alongside other common and rare disease datasets.

**NHGRI’s Genomic Data Science Analysis, Visualization, and Informatics Lab-space (AnVIL)** is a cloud-based environment where both data and tools can co-exist, thereby improving logistics for the wider community to be able to share and access them together. AnVIL is assembling the most commonly-used tools and pipelines to support genomic analysis and make them available on the AnVIL platform. In addition, investigators can add their own tools to the platform. There are currently a number of workflows important in rare disease analysis set up in AnVIL, including germline variant calling with GATK, RNAseq processing, and mitochondrial variant calling. The *seqr* analysis platform is on AnVIL and any researcher can request that a jointly-called vcf located within an AnVIL workspace be loaded in *seqr*. By creating an environment to share tools and pipelines, the application of analysis methods to datasets will be facilitated, allowing for comparison of the performance of approaches to each other to help develop best practices. The introduction of novel tools and methods will be immediately leveraged and readily testable. This will ultimately allow authorized researchers anywhere in the world to explore their own hypotheses using CMG data and a constantly evolving set of analysis tools.

While the tools and workflows are open access for anyone logging into the interface, AnVIL datasets have three types of data access: open access, controlled-access, and consortium access. To learn more about the types of AnVIL access and examples, see Table 4. All CMG datasets are controlled-access, meaning researchers must request access through dbGaP and obtain permission for use consistent with the subjects’ informed consent (e.g. General Research Use (GRU), Health/medical/biomedical research and clinical care (HMB), Research use only (RUO)). Access is regulated in accordance with the NIH policy and full details can be found at https://anvilproject.org/data/requesting-data-access.

**Table 4.** Access Levels in AnVIL

| **Access type** | **Description** | **Example Dataset** | **Additional documentation** |
| --- | --- | --- | --- |
| Open Access | These data sets and tools are accessible to anyone with an AnVIL, Terra or Gen3 account. | 1000 Genomes | https://anvilproject.org/learn |
| Controlled-Access | Access to these data sets is managed by a data access committee designated by dbGaP and is based on intended use of the requester and allowed use of the data submitter as defined by consent codes. | CMG cohorts, GTEx | https://anvilproject.org/learn/accessing-data/requesting-data-access#accessing-controlled-access-data |
| Consortium Access | These data sets are accessible to consortia members and controlled under the consortium's data sharing agreement. | Centers for Common Disease Genomics (CCDG), Cohorts that are not yet released from their embargo period | https://anvilproject.org/learn/for-consortia/consortium-data-access-guidelines |

As of this publication, the CMGs have deposited over 15,025 exomes and 707 genomes to 39 AnVIL workspaces (https://anvilproject.org/data). In addition to the raw sequencing data, the CMGs have uploaded the accompanying metadata for each sample, including sample-, subject-, family-, discovery- and sequence-level information (see supplemental table 3 for file formats). The subject metafile includes description of the individual including but not limited to ancestry, sex, relationship of family members, and phenotype description. The subject metafile provides sample-level details including source of sample type and sequencing center, while the family metafile addresses consanguinity and pedigree information. The discovery metafile includes both gene- and variant-level details for known and candidate genes for potentially solved cases. Finally, the sequencing metafile provides context for the sequencing files including sequencing metrics, file paths, and assay details. The data model addresses monogenic disease due to small DNA variation or large copy number variation, as well as special use cases such as multigenic cases and disorders caused by somatic variation.

### Recommendations for accessing CMG data through AnVIL

The CMG Program-sequenced data in AnVIL contains pathogenic variants in genes with both known and novel gene-disease associations, which have not been identified by the analysis strategies taken to date. These variants may not have been identified due to differing variant/gene prioritization approaches, data parsing computational approaches, variant filtering parameters implemented in computational pipelines, or may not have been robustly detected or optimized by existing calling methods at the time of analysis. Thus, we envision that primary uses of CMG data in AnVIL will be for novel gene-disease discovery and for identification of pathogenic variants previously not detected or able to be interpreted. Aggregation across all CMG datasets could enable large-scale gene discovery via association or enrichment/burden analyses of rare disease phenotypes for which data from many affected individuals are available, i.e. “intellectual disability,” “epilepsy,” “brain malformations.” Similarly, application of a broad range of variant calling methods could enable curation of a benchmark set of variants for use by future developers of callers for structural variants, repeat expansions, and other “challenging” variant classes.

We urge future users to consider potential caveats to the data released in AnVIL. Users should be aware that samples may have variable quality (e.g. exceptionally valuable samples may have been sequenced even though DNA quality was not ideal) and that some of the data being shared was generated in the “early days” of exome sequencing. Data in AnVIL have not been reprocessed and recalled using a single pipeline, so users should anticipate variability in variant aligners, callers, and calling targets, in addition to the prior choices of different exome capture methods. Finally, deep phenotypic information is not accessible or available for some individuals due to a variety of reasons (e.g. lack of funding for deep phenotyping, loss of an individual to follow-up studies, etc.). Future users may need to contact the CMG to be connected with the original submitters of a dataset to seek additional phenotype information and/or collaborate to collect such data.

### Data sharing empowers and expedites solving rare disease

There are a number of unsolved syndromes that have perplexed the clinical genetics community for decades.^40^ Although collectively the CMGs have sequenced over 28,991 families, each individual CMG often has only a handful of cases of a given phenotype.

We formed the CMG Data Analysis working group to share cohorts across CMGs in order to increase power to solving these challenging phenotypes. For our pilot project, we gathered all cases with a clinical diagnosis of Dubowitz syndrome, and reached out to CMG collaborators to gather additional cases. In total, we were able to build a cohort of 20 individuals from 16 families. It would have been very challenging to collect this many families with a rare condition without access to an international network of researchers and clinicians. The University of Washington CMG reprocessed the data through a GATK pipeline and generated a joint-called vcf. Upon analysis, no two cases shared the same genetic diagnosis and no specific pathways were identified. A collaboration was also established with the Canadian Care4Rare Program, which had been building a similar cohort. Only after combining the CMG and FORGE/Care4Rare cohorts (31 individuals from 27 families) was the group able to recognize that the diagnosis of Dubowitz syndrome was not pointing to a single disorder but a collection of disorders with overlapping phenotypic features, highlighting the benefit of data aggregation for these studies. Overall, we found that the Dubowitz syndrome phenotype has extensive locus heterogeneity rather than a single gene disorder. Diagnoses were made for a number of recently molecularly defined and phenotypically similar conditions with growth restriction, microcephaly and developmental delay, with a molecular diagnosis made in 13/27 families (48%) or strong candidate variants in known and candidate disease genes identified in an additional 7/27 families (26%).^41^ Only one gene*, HDAC8,* was linked to the phenotype in multiple families (2 families). This experience highlights the need to continue to work across national and international rare disease programs to build larger cohorts for rare conditions.

Model organism data are an important component of gene discovery. Even when multiple families with overlapping phenotypes have had a variant identified in the same gene, establishing a causal gene-disease relationship can be difficult. As part of the effort to clarify these relationships, the CMGs formed a collaboration with the Knock-Out Mouse Project (KOMP) whose mission was to knock out every mouse gene and perform detailed phenotyping of null alleles (https://www.komp.org). Given the resource limitations of this program, it was important to prioritize genes that may be important for human disease. We shared all candidate genes after their respective embargo period (tier 1, 13 months; tier 2, 19 months) or earlier at collaborator request. CMG collaborators could sign up to track the gene within the International Mouse Phenotype Consortium (IMPC) database and request to purchase the mouse once it was generated. Several gene discoveries including *TONSL* skeletal dysplasia and *FAM92A* limb and digit anomalies were supported by this collaborative mechanism.^42, 43^

### Development of tools and improved methods

The CMGs have developed the open source analysis tools PhenoDB and *seqr* for filtering and prioritizing variants in individuals or families with Mendelian disease.^44, 45^ These platforms enable streamlined analysis of exome or genome sequence data through incorporation of *in silico* predictions, population and disease databases, as well as integration with other external databases to facilitate review of novel candidates (e.g. GeneCards, Mouse Genome Informatics). Clinical and pedigree data can also be recorded, including in a standardized format (e.g. Human Phenotype Ontology), enabling coupling of genetic and phenotypic data. These web-based tools facilitate collaboration by providing a platform for researchers from disparate locations to work as a team on analyses, including by enabling tagging and adding notes to variants of interest and by recording the analysis status of each case (i.e. solved, in progress). By updating in real time, these tools avoid version control issues while facilitating easy tracking of diagnostic yields across cohorts. Furthermore, these tools offer the ability to directly submit genes of interest to the MME. PhenoDB and *seqr* continue to be periodically updated and are available for free download and implementation, thereby providing broadly-useful resources for Mendelian disease research.

The CMGs have also developed or contributed to methods to reanalyze exome data from unsolved cases. This includes semi-automated pipelines for periodic reanalysis, where analysis of exome and phenotype data with updated methods and annotations yielded confirmed or potential genetic diagnoses in up to a third of unsolved cases, and mostly within disease genes published after initial analysis.^46, 47^ While the pace of disease gene discovery emphasizes the importance of periodic reanalysis, it is the automation of these processes that will meet the challenge of a continued accrual of unsolved cases. CMG investigators have also developed and applied ‘gene-centric’ analyses to identify candidate disease genes in exome-negative cohorts, such as burden tests, to identify genes enriched in deleterious rare variants across cases with the same phenotype,^48, 49^ and a phenotype-agnostic method that prioritizes genes most likely to underlie Mendelian disease.^50^ Reanalysis methods for more complicated patterns of inheritance led to the development of a two-locus genome-wide test that enables detection of digenic inheritance in exome data.^51^

CMGs investigators have successfully applied sequencing approaches beyond the exome to identify or validate causal variant(s), including genome, RNA, bisulfite DNA methylation sequencing, and long-read sequencing, showing their utility in cases where exome sequencing fails to find a molecular diagnosis. Examples include utilizing genome sequencing to identify pathogenic SVs missed by exome, such as the homozygous inversion in *QDPR* detected in a patient with dihydropteridine reductase deficiency;^52^ applying RNA sequencing to identify genes with aberrant expression and or splicing, including an intronic variant *in trans* with a missense in muscle disease gene *DES* that resulted in a pseudo-exon insertion and allelic imbalance;^53^ using bisulfite sequencing to identify gene silencing epivariation, such as the characterization of aberrant hypermethylation associated with a pathogenic repeat expansion in the *XYLT1* promoter region;^54^ and the application of long-read sequencing to characterize a complex genomic rearrangement involving an inverted triplication flanked by duplications in a proband with Temple syndrome.^55^

The challenges with interpreting rare variation in the genome is a major barrier for achieving genetic diagnoses. To this end, CMGs investigators have built resources and tools to improve variant interpretation. Analysis of the Genome Aggregation Database (gnomAD) provided insight into the distribution of structural, multi-nucleotide, and untranslated region variants in reference populations, enabling improved interpretation of these often difficult to annotate variants.^56–58^ Updated pseudogene annotation for the human reference genome, and improved annotation of enhancers and promoter regions, both achieved by leveraging data from the ENCODE project, can further facilitate variant interpretation, particularly for genome analyses.^59, 60^ In particular, tissue-specific genomic annotations generated as part of the EN-TEX project will play a key role in interpreting non-coding variants in the genome.^61^ The GTEx consortium transcriptomics data facilitated the development of NMDEscPredictor and whether a PTC may ‘escape’ nonsense mediated decay.^62^ Similarly, there has been a recent effort toward identifying pathogenic structural variations in various rare and common diseases while taking into account of the tissue-specific epigenomic, genomics, and conservation-related features.^63^

## Discussion

As the CMG program comes to a close, additional work is still needed to identify the genetic basis of Mendelian disease for many families. Hundreds of novel gene-disease candidates were identified but many still lack sufficient data to confirm or refute the relationship. Over half of the families sequenced remain unsolved despite having phenotypes that are strongly suggestive of a Mendelian cause. There are also thousands of genes predicted by human or model organism data to result in a human phenotype when disrupted that have not yet been linked to a human phenotype.^64, 65^ The mission started by the CMGs remains far from finished, but the data and tools developed will continue to support the ongoing research efforts in this area.

New bioinformatic methods and resources are needed to help address the limitations of current analytical approaches and data (ex., exomes vs genomes, short read vs long read tech, limited pheno data). The telomere to telomere genome assembly and the use of a ‘pan-genome’ human reference sequence representing diverse ancestries will improve variant interpretation;^66, 67^ however, it will also require new analytical pipelines,^68^ including support for graph-based representation typically used for pan-genome data.^69^ Methods for evaluating cases with incomplete penetrance and digenic or oligogenic inheritance are also needed.

Efforts to resolve VUS, particularly missense variant alleles, are needed. With progress in recent years for deep mutational scanning, there is now an international focus through the Atlas of Variant Effect Alliance, which aims to systematically determine the impact of variants in functionally important genomic regions.^70^ The application of artificial intelligence methods for predicting the functional effect of variants also offers an avenue for improving *in silico* predictions of pathogenicity.^71, 72^ Much of our knowledge about gene and transcript expression patterns relies on adult tissue; the Developmental Genotype-Tissue Expression (dGTEx) and Pediatric Cell Atlas initiatives will facilitate interpretation for developmental disorders.^73^

Population datasets and analytical tools that aid the interpretation of long-read sequencing and methylation data in rare disease patients will be needed to help realize the diagnostic utility of these technologies. Furthermore, the integration of methylation, structural and RNA-seq data into variant analytical tools will ultimately be required to streamline analysis for cases remaining unsolved after exome sequencing, particularly for compound heterozygotes where only one of the variants will be detected and prioritized by exome or genome.

While the CMGs strived to include collaborators and cases from around the world (Figure 3), there is still progress to be made on the diversity of both the participating researchers we collaborate with and the genetic ancestry of the cases we sequence. Seventy three percent of collaborators reported their ancestry to be White/European, and the majority (58%) were male. Additionally, 66.75% percent of the cases sequenced were also White/European. To make good on the promise of genomics, a much greater fraction of the world’s populations must be involved. Research programs must push to expand the diversity of the research workforce as well as the families and individuals studied. One challenge has been that cases from countries with less access to genomic research and medicine likely lack prior genetic testing, yielding a higher number of diagnoses in known disease genes that would be identified by routine testing.

**Figure 3.**
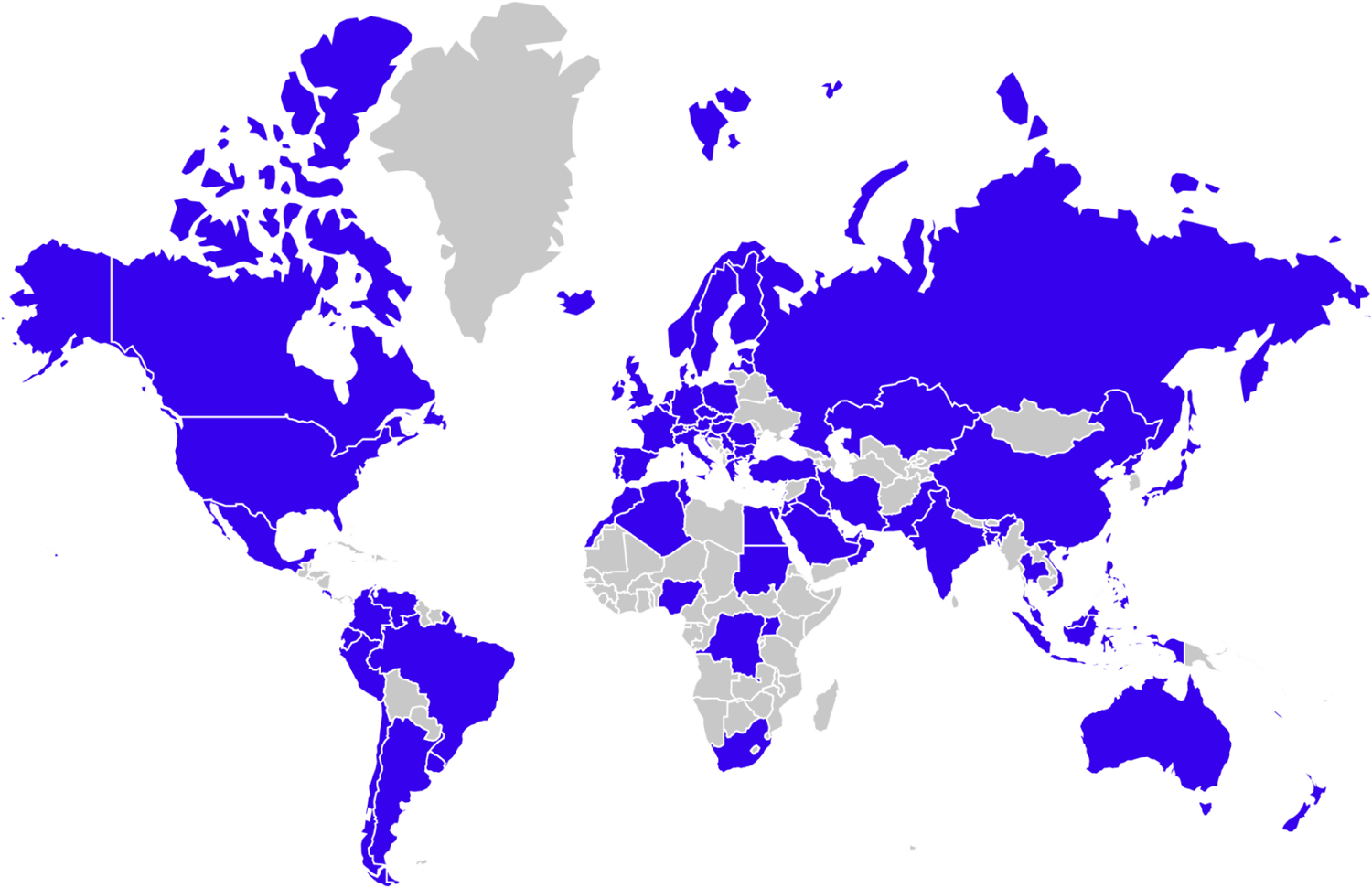
Map of CMG collaborators. Blue indicates that the CMGs collaborated with at least one researcher in that country (based on country listing in pubmed affiliations).

These cases might be best approached by higher throughput clinical-research partnerships with philanthropic or other sources of funding followed by data sharing of negative cases with research centers able to apply advanced techniques.

Debate continues regarding models for funding rare disease research, either distributing funds to individual rare disease investigators working in their own fields versus centralizing funding for centers that can build infrastructure used by many investigators. The success of the CMG collaborators in discoveries, publications, data sharing, and subsequent research funding highlights the power of centralized funding for centers.

The CMGs structure has allowed for sequencing and analysis of invaluable samples to be performed in centers of excellence by a team of experts and more than that, it has facilitated investigators of the same rare disease around the world to connect and collaborate. This approach offers cost efficiency by distributing shared infrastructure across large numbers of investigators as well as enabling better data sharing and cohort aggregation for increased statistical power. In this decade of gene discovery by the CMGs, there has been substantial progress made but much remains to be done. As routine genomic analysis becomes more successful, approaching a 50% diagnosis rate in clinical diagnostic labs, the cases that remain unsolved are increasingly challenging. Undiscovered diseases are ultra-rare, the functional impact of variation is difficult to determine, and causality is hard to prove, particularly for variants with incomplete penetrance and complex genetic architectures. Gene discovery rates remain steady, highlighting the continued need for national and international programs in rare disease genomic analysis, including the recently funded Mendelian Genomics Research Consortium (https://www.genome.gov/news/news-release/NIH-funds-new-effort-to-discover-genetic-causes-of-single-gene-disorders).

## Data Availability statement

Candidate genes identified by the CMG have been submitted to Matchmaker Exchange (https://www.matchmakerexchange.org). Variant classifications have been submitted to ClinVar. De-identified and coded genomic and phenotype data have been shared on the National Human Genome Research Institute (NHGRI) AnVIL platform. Data access requests can be made per instructions here https://anvilproject.org/learn/accessing-data/requesting-data-access#accessing-controlled-access-data.

## Data Availability

Candidate genes identified by the CMG have been submitted to Matchmaker Exchange (https://www.matchmakerexchange.org/). Variant classifications have been submitted to ClinVar. De-identified genomic and phenotype data have been shared on the National Human Genome Research Institute (NHGRI) AnVIL platform. Data access requests can be made per instructions here https://anvilproject.org/learn/accessing-data/requesting-data-access#accessing-controlled-access-data.

https://anvilproject.org/data?query=consortium%3DCMG

https://www.ncbi.nlm.nih.gov/clinvar/submitters/505572/

https://www.ncbi.nlm.nih.gov/clinvar/submitters/505755/

https://www.ncbi.nlm.nih.gov/clinvar/submitters/506627/

https://www.ncbi.nlm.nih.gov/clinvar/submitters/505516/

https://www.ncbi.nlm.nih.gov/clinvar/submitters/506150/

http://mendelian.org/phenotypes-genes

## Acknowledgements

The Baylor Hopkins Center for Mendelian Genomics, Broad Institute Harvard Center for Mendelian Genomics, University of Washington Center for Mendelian Genomics, and Yale Center for Mendelian Genomics were funded by the National Human Genome Research Institute (NHGRI) awards UM1HG006542, UM1HG008900, UM1HG006493, and UM1HG006504, respectively. Funds were also provided under the National Heart, Lung, and Blood Institute (NHLBI) under the Trans-Omics for Precision Medicine Program (TOPMed), and the National Eye Institute (NEI). The GSP Coordinating Center (U24HG008956) contributed to cross-program scientific initiatives and provided logistical and general study coordination. Aspects of this work were funded by NHGRI K08HG008986 (JEP), NHGRI R01HG009141 (HLR and DGM), the National Institute of Neurological Disorders and Stroke (NINDS) R35NS105078 (JRL), and National Health and Medical Research Council (NHMRC) Early Career Fellowship 1159456 (NJL). The CMGs would like to thank all of our collaborators from around the world as well as the families and individuals who contributed their data to this study.

## Author Contributions

Conceptualization: S.M.B., J.E.P, N.J.L, N.S., J.X.C., S.B., E.E.B, L.H.C., K.F.D., C.P.D., M.L. C.W., S.N.J., R.A.G., R.P.L., D.G.M., M.G., T.C.M., J.R.L., D.V., M.J.B., A.H., S.M., D.A. N., H.L.R., A.O.L.; Data curation: S.M.B., J.E.P, N.J.L, N.S., J.X.C., S.B., A.H., K.F.D; Formal analysis: S.M.B., J.E.P, N.J.L, N.S., J.X.C., S.B., Z.H.C., S.N.J., M.J.B., A.H., S.M., D.A. N., H.L.R., A.O.L..; Funding acquisition: S.B., R.A.G., R.P.L., D.G.M., M.G., T.C.M., J.R.L., D.V., M.J.B., A.H., S.M., D.A.N., H.L.R., K.F.D.; Project administration: L.H.C., C.W.; Visualization: S.M.B., S.B., A.O.L.; Writing–original draft: S.M.B., J.E.P, N.J.L, N.S., J.X.C., S.B., M.L., M.J.B., A.H., S.M., H.L.R., A.O.L.; Writing–review & editing: S.M.B., J.E.P, N.J.L, N.S., J.X.C., S.B., E.E.B, L.H.C., K.F.D., C.P.D., M.L., Z.H.C., C.W., S.N.J., R.A.G., R.P.L., D.G.M., M.G., T.C.M., J.R.L., D.V., M.J.B., A.H., S.M., D.A. N., H.L.R., A.O.L.

## Ethics Declaration

Informed consent was obtained by collaborators for all participants in studies across the CMGs, and individual-level data, including genomics and clinical data, was de-identified and coded by our collaborators prior to submission to the CMGs. The participant’s samples used for this study were obtained from multiple institutions and each CMG (Baylor-Hopkins, Broad Institute of MIT and Harvard, University of Washington, Yale) was responsible for submitting to their own IRB to receive local approval.

## Supplemental Table Captions

Supplemental Table 1. CMG discovery case summary sheet

Supplemental table 2. CMG data sharing details

Supplemental Table 3. CMG AnVIL metadata dictionary and example files

## Centers for Mendelian Genomics Consortium

Marcia Adams^1^, François Aguet^2^, Gulsen Akay^3^, Peter Anderson^4^, Corina Antonescu^1^, Harindra M. Arachchi^5^, Mehmed M. Atik^3^, Christina A. Austin-Tse^5,6^, Larry Babb^5^, Tamara J. Bacus^4^, Vahid Bahrambeigi^3^, Suganthi Balasubramanian^7,8^, Yavuz Bayram^3^, Arthur L. Beaudet^3^, Christine R. Beck^3^, John W. Belmont^3^, Jennifer E. Below^9^, Kaya Bilguvar^10^, Corinne D. Boehm^1^, Eric Boerwinkle^9,11^, Philip M. Boone^3^, Sara J. Bowne^9^, Harrison Brand^12^, Kati J. Buckingham^4^, Alicia B. Byrne^5,13^, Daniel Calame^3^, Ian M. Campbell^3^, Xiaolong Cao^14^, Claudia M.B. Carvalho^3^, Varuna Chander^11^, Jaime Chang^5^, Katherine R. Chao^5^, Ivan K. Chinn^15,16^, Declan Clarke^7,17^, Ryan L. Collins^6,12,18^, Beryl Cummings^5^, Zain Dardas^3^, Moez Dawood^3^, Kayla Delano^5^, Stephanie P. DiTroia^5^, Harshavardhan Doddapaneni^11^, Haowei Du^3^, Renqian Du^3^, Ruizhi Duan^3^, Mohammad Eldomery^3^, Christine M. Eng^3,19^, Eleina England^5^, Emily Evangelista^5^, Selin Everett^5^, Jawid Fatih^3^, Adam Felsenfeld^20^, Laurent C. Francioli^5^, Christian D. Frazar^4^, Jack Fu^12^, Emmanuel Gamarra^14^, Tomasz Gambin^3^, Weiniu Gan^21^, Mira Gandhi^3^, Vijay S. Ganesh^5,22,23^, Kiran V. Garimella^24^, Laura D. Gauthier^5^, Danielle Giroux^4^, Claudia Gonzaga-Jauregui^3^, Julia K. Goodrich^5^, William W. Gordon^4^, Sean Griffith^1^, Christopher M. Grochowski^3^, Shen Gu^3^, Sanna Gudmundsson^5,23^, Stacey J. Hall^5^, Adam Hansen^11^, Tamar Harel^3,25^, Arif O. Harmanci^17,26^, Isabella Herman^3^, Kurt Hetrick^1^, Hadia Hijazi^3^, Martha Horike-Pyne^4^, Elvin Hsu^1^, Jianhong Hu^11^, Yongqing Huang^24^, Jameson R. Hurless^4^, Steve Jahl^5^, Gail P. Jarvik^4^, Yunyun Jiang^11^, Eric Johanson^4^, Angad Jolly^3^, Ender Karaca^3^, Michael Khayat^11^, James Knight^10^, J. Thomas Kolar^4^, Sushant Kumar^7^, Seema Lalani^3,19^, Kristen M. Laricchia^5^, Kathryn E. Larkin^5^, Suzanne M. Leal^27^, Gabrielle Lemire^5^, Richard A. Lewis^3^, He Li^11^, Hua Ling^1^, Rachel B. Lipson^5^, Pengfei Liu^3,19^, Alysia Kern Lovgren^5^, Francesc López-Giráldez^10^, Melissa P. MacMillan^4^, Brian E. Mangilog^5^, Stacy Mano^5^, Dana Marafi^3,11,15^, Beth Marosy^1^, Jamie L. Marshall^5^, Renan Martin^1^, Colby T. Marvin^4^, Michelle Mawhinney^1^, Sean McGee^4^, Daniel J. McGoldrick^4^, Michelle Mehaffey^28^, Betselote Mekonnen^4^, Xiaolu Meng^3^, Tadahiro Mitani^3^, Christina Y. Miyake^15,29^, David Mohr^1^, Shaine Morris^15,29^, Thomas E. Mullen^5^, David R. Murdock^3,11^, Mullai Murugan^11^, Donna M. Muzny^11^, Ben Myers^1^, Juanita Neira^3^, Kevin K. Nguyen^5^, Patrick M. Nielsen^4^, Natalie Nudelman^14^, Emily O’Heir^5^, Melanie C. O’Leary^5^, Chrissie Ongaco^1^, Jordan Orange^15,30,31^, Ikeoluwa A. Osei-Owusu^5^, Ingrid S. Paine^3^, Lynn S. Pais^5^, Justin Paschall^1^, Karynne Patterson^4^, Davut Pehlivan^32^, Benjamin Pelle^4^, Samantha Penney^3^, Jorge Perez de Acha Chavez^5^, Emma Pierce-Hoffman^5^, Cecilia M. Poli^33,34^, Jaya Punetha^3^, Aparna Radhakrishnan^4^, Matthew A. Richardson^4^, Eliete Rodrigues^1^, Gwendolin T. Roote^4^, Jill A. Rosenfeld^3^, Erica L. Ryke^4^, Aniko Sabo^11^, Alice Sanchez^1^, Isabelle Schrauwen^27^, Daryl A. Scott^3^, Fritz Sedlazeck^11^, Jillian Serrano^5^, Chad A. Shaw^3,19^, Tameka Shelford^1^, Kathryn M. Shively^4^, Moriel Singer-Berk^5^, Joshua D. Smith^4^, Hana Snow^5^, Grace Snyder^20^, Matthew Solomonson^5^, Rachel G. Son^5^, Xiaofei Song^3^, Pawel Stankiewicz^3,19^, Taylorlyn Stephan^20,35^, V. Reid Sutton^3,16^, Abigail Sveden^5^, Diana Cornejo Sánchez^27^, Monica Tackett^4^, Michael Talkowski^12^, Machiko S. Threlkeld^4^, Grace Tiao^5^, Miriam S. Udler^5,6^, Laura Vail^1^, Zaheer Valivullah^5^, Elise Valkanas^5^, Grace E. VanNoy^5^, Qingbo S. Wang^5^, Gao Wang^27^, Lu Wang^36^, Michael F. Wangler^3^, Nicholas A. Watts^5^, Ben Weisburd^5^, Jeffrey M. Weiss^4^, Marsha M. Wheeler^4^, Janson J. White^3^, Clara E. Williamson^5^, Michael W. Wilson^5^, Wojciech Wiszniewski^3^, Marjorie A. Withers^3^, Dane Witmer^1^, Lauren Witzgall^5^, Elizabeth Wohler^1^, Monica H. Wojcik^5,37^, Isaac Wong^12^, Jordan C. Wood^5^, Nan Wu^3,38^, Jinchuan Xing^14^, Yaping Yang^3,19^, Qian Yi^4^, Bo Yuan^3,39,40^, Jordan E. Zeiger^4^, Chaofan Zhang^3^, Peng Zhang^1^, Yan Zhang^1^, Xiaohong Zhang^4^, Yeting Zhang^14^, Shifa Zhang^5^, Huda Zoghbi^3^, Igna van den Veyver^3^

^1^McKusick-Nathans Department of Genetic Medicine, Johns Hopkins University School of Medicine, Baltimore, MD, USA

^2^Broad Institute of MIT and Harvard, Cambridge, MA, USA

^3^Department of Molecular and Human Genetics, Baylor College of Medicine, Houston, TX, USA

^4^Division of Genetic Medicine, Department of Pediatrics, University of Washington, Seattle, WA, USA

^5^Broad Center for Mendelian Genomics, Broad Institute of MIT and Harvard, Cambridge, MA, USA

^6^Center for Genomic Medicine, Massachusetts General Hospital, Boston, MA, USA

^7^Department of Molecular Biophysics and Biochemistry, Yale University, New Haven, CT, USA

^8^Regeneron Genetics Center, Regeneron Pharmaceuticals, Tarrytown, NY, USA

^9^Human Genetics Center, University of Texas Health Science Center at Houston School of Public Health, Houston, TX, USA

^10^Yale Center for Genome Analysis, Yale University, New Haven, USA

^11^Human Genome Sequencing Center, Baylor College of Medicine, Houston, TX, USA

^12^Program in Medical and Population Genetics, Broad Institute of MIT and Harvard, Cambridge, MA, USA

^13^UniSA Clinical and Health Sciences, University of South Australia, Adelaide, SA, Australia

^14^Department of Genetics, Rutgers University, Piscataway, NJ, USA

^15^Department of Pediatrics, Baylor College of Medicine, Houston, TX, USA

^16^Department of Pediatrics, Texas Children’s Hospital, Houston, TX, USA

^17^Program in Computational Biology and Bioinformatics, Yale University, New Haven, CT, USA

^18^Program in Bioinformatics and Integrative Genomics, Harvard Medical School, Boston, MA, USA

^19^Baylor Genetics diagnostic laboratory, Houston, TX, USA

^20^Division of Genome Sciences, National Human Genome Research Institute, Bethesda, MD, USA

^21^Division of Lung Diseases, National Heart, Lung and Blood Institute, Bethesda, MD, USA

^22^Neuromuscular Division, Department of Neurology, Brigham and Women’s Hospital, Boston, MA, USA

^23^Division of Genetics and Genomics, Boston Children’s Hospital, Boston, MA, USA

^24^Data Science Platform, Broad Institute of MIT and Harvard, Cambridge, MA, USA

^25^Department of Genetic and Metabolic Diseases, Hadassah-Hebrew University Medical Center, Jerusalem, Israel

^26^Center for Precision Medicine, School of Biomedical Informatics, University of Texas Health Science Center Houston, Houston, TX, USA

^27^Center for Statistical Genetics, Columbia University, New York, NY, USA

^28^Pacific Northwest Research Institute, Seattle, WA, USA

^29^Department of Pediatrics, Division of Cardiology, Texas Children’s Hospital, Houston, TX, USA

^30^Department of Pediatrics, Section of Immunology, Allergy and Rheumatology, Texas Children’s Hospital, Houston, TX, USA

^31^Department of Pediatrics, Vagelos College of Physicians and Surgeons, Columbia University Irving Medical Center, New York, NY, USA

^32^Department of Neurology, Texas Children’s Hospital, Houston, TX, USA

^33^Department of Pediatrics, Section of Immunology, Allergy and Rheumatology, Baylor College of Medicine, Houston, TX, USA

^34^Clinica Alemana de Santiago, Facultad de Medicina Clinica Alemana-Universidad del Desarrollo, Santiago, Chile

^35^Department of Biomolecular Engineering and Bioinformatics, University of California, Santa Cruz, Santa Cruz, CA, USA

^36^Translational Genomics Research Branch, National Institute of Dental and Craniofacial Research, Bethesda, MD, USA

^37^Divisions of Newborn Medicine and Genetics and Genomics, Boston Children’s Hospital, Boston, MA, USA

^38^Department of Orthopedic Surgery, Peking Union Medical College and Chinese Academy of Medical Sciences, Beijing, China

^39^Seattle Children’s Hospital, Seattle, WA, USA

^40^Department of Laboratory Medicine and Pathology, University of Washington, Seattle, WA, USA

## Consortium members received funding as follows

Alicia B. Byrne: A.B.B. was supported by the Australian Government Research Training Program Scholarship, the Australian Genomics Health Alliance & NHMRC (GNT1113531) and the Maurice de Rohan International Scholarship Laurent C. Francioli: L.C.F. was supported by the Swiss National Science Foundation (Advanced Postdoc.Mobility 177853) Vijay S. Ganesh: V.S.G. was supported by NIH/NHGRI T32HG010464 Sanna Gudmundsson: S.G. was supported by the Knut and Alice Wallenberg Foundation Miriam S. Udler: MSU was supported by NIH/NIDDK K23DK114551 Monica H. Wojcik: MHW was supported by NIH/NICHD K23HD102589 and by an Early Career Award from the Thrasher Research Fund

