## Supplemental Table 2 for "Centers for Mendelian Genomics: A decade of facilitating gene discovery"

**Supplemental table 2: CMG data sharing details**

| Center | ClinVar Name | ClinVar URL | dbGaP/AnVIL Study accession ID |
|---|---|---|---|
| Baylor-Hopkins | Lupski Lab, Baylor-Hopkins CMG, BHCMG | https://www.ncbi.nlm.nih.gov/clinvar/submitters/505572/ | phs000711 |
| Baylor-Hopkins | Baylor-Hopkins Center for Mendelian Genomics (Johns Hopkins University) | https://www.ncbi.nlm.nih.gov/clinvar/submitters/505755/ | phs000711 |
| Broad Institute | Broad Institute Rare Disease Group (Broad Institute) | https://www.ncbi.nlm.nih.gov/clinvar/submitters/506627/ | phs001272 |
| University of Washington | University of Washington Center for Mendelian Genomics (University of Washington), UW-CMG | https://www.ncbi.nlm.nih.gov/clinvar/submitters/505516/ | phs000693 |
| Yale | Yale Center for Mendelian Genomics (Yale University) | https://www.ncbi.nlm.nih.gov/clinvar/submitters/506150/ | phs000744 |
